# Hearing loss is associated with gray matter differences in older adults at risk for and with Alzheimer’s disease

**DOI:** 10.1101/2020.10.07.20208017

**Authors:** N. Giroud, M. K. Pichora-Fuller, P. Mick, W. Wittich, F. Al-Yawer, S. Rehan, J.B. Orange, N. A. Phillips

## Abstract

Using data from the COMPASS-ND study we investigated associations between hearing loss and hippocampal volume as well as cortical thickness in older adults with subjective cognitive decline (SCD), mild cognitive impairment (MCI), and Alzheimer’s dementia (AD). SCD participants with greater pure-tone HL exhibited lower hippocampal volume, but more cortical thickness in the left superior temporal gyrus and right pars opercularis. Greater speech-in-noise reception thresholds were associated with lower cortical thickness bilaterally across much of the cortex in AD. The AD group also showed a trend towards worse speech-in-noise thresholds compared to the SCD group.

**Highlights:**

- In SCD, greater pure-tone hearing loss was associated with lower right hippocampal volume.
- Pure-tone hearing loss was not associated with brain atrophy in MCI or AD.
- Individuals with AD exhibited a trend towards poorer speech-in-noise (SiN) thresholds than SCD.
- In AD, greater atrophy across large portions of the cortex was associated with greater SiN thresholds.

## 1. Introduction

The global prevalence of Alzheimer’s dementia (AD) is expected to triple by 2050, leading to immense personal, social, and health care costs^1^. Attention is now focused on behavioral and non-pharmacological interventions because of the low efficacy of pharmacological treatments (Livingston et al., 2017, 2020). Identifying and treating modifiable risk factors is a promising strategy to delay the onset or progression of AD (Livingston et al., 2017, 2020). Indeed, it is estimated that delaying the onset of dementia by 5 years would lead to an approximate 50% reduction in prevalence after 10 years (Brookmeyer et al., 1998). In a meta-analysis by Livingston et al. (2020) examining risk factors for dementia, the population attributable fraction for hearing loss was estimated at 8%, which was higher than the value for all other modifiable risk factors identified in the study. Currently, there is insufficient evidence regarding whether or not prevention of or treatments for hearing loss can modify dementia risk, but this is an active topic of research (Deal et al., 2018; Sanchez et al., 2020; Sarant et al., 2020).

Typically, age-related hearing loss is characterized by elevated pure-tone audiometric thresholds for high-frequency sounds (International Organization for Standardization (ISO), 2017). Age-related hearing loss is multifactorial, with increased audiometric thresholds at high-frequencies often resulting from damage to cochlear outer hair cells or the stria vascularis in the auditory periphery (Dubno et al., 2013; Mills et al., 2006). In addition, there is evidence that degeneration in the synaptic connections between cochlear hair cells and nerve fibers may contribute to inaccurate coding of acoustic signals, leading to difficulties speech understanding (Liberman and Kujawa, 2017). Peripheral auditory damage can also lead to maladaptive (sub)cortical plasticity leading, for example, to a loss of inhibition and hyperexcitability (Herrmann and Butler, 2020). Notably, even older adults with normal or near-normal audiometric pure-tone thresholds can have difficulties understanding speech in noise in everyday situations because of age-related declines in auditory processing (Pichora-Fuller et al., 2017). In general, age-related hearing loss leads to difficulty participating in conversations and social interactions and is associated with reduced quality of life, social isolation, and higher rates of depressive symptoms (Arlinger, 2003; Pichora-Fuller et al., 2015). Older adults often remark that they can hear but cannot discriminate or easily understand what is said in noisy environments.

There is a strong link between auditory and cognitive functioning (e.g. Lindenberger and Baltes, 1994). Hearing loss (defined by pure-tone thresholds or measures of auditory processing such as speech-in-noise understanding) is related to self-reported (Curhan et al., 2019) and behavioral measures of cognitive decline in aging (Fischer et al., 2016; Fortunato et al., 2016; de la Fuente et al., 2019; Merten et al., 2019). Hearing loss also is linked to incident all-cause dementia (Albers et al., 2015; Deal et al., 2015, 2019; Gates et al., 2011; Lin and Albert, 2014; Lin et al., 2011a; Osler et al., 2019). However, despite growing evidence for a link between auditory and cognitive decline, the underlying mechanisms are still unclear. Several hypotheses were proposed over 25 years ago to explain the relationship between hearing and cognitive decline (Lindenberger and Baltes, 1994, and versions of these hypotheses continue to motivate research; e.g., Whitson et al., 2018); however, no single hypothesis can explain most of the effects reported in the literature (Pronk et al., 2019). Consistent with the common-cause hypothesis, associations between hearing and cognitive decline may be due to a common biological cause such as widespread age-related neural decline. According to the information degradation hypothesis, it is possible that older adults do not encode auditory information as well as those with normal hearing. When it is difficult to hear, such as in noisy environments, the listener may increase the allocation of cognitive resources to lower-level perceptual auditory processing, thereby diverting resources from higher-order cognitive processing and resulting in poorer cognitive performance (e.g., on measures of memory (McCoy et al., 2005)). In addition, according to the sensory deprivation hypothesis, chronic reallocation of cognitive resources may bring about permanent changes in patterns of brain activation and structure (Peelle and Wingfield, 2016).

In addition to the auditory-cognitive link, an association between hearing loss and brain atrophy in gray and white matter has been reported. Greater hearing loss is correlated with lower gray matter volume in brain regions associated with auditory perception (e.g., superior temporal lobe) as well as regions associated with cognition (e.g., hippocampus, parahippocampus) (Alfandari et al., 2018; Armstrong et al., 2019; Eckert et al., 2012; Ren et al., 2018; Rigters et al., 2018, 2017; Rudner et al., 2019; Tuwaig et al., 2017; Uchida et al., 2018; but see Profant et al., 2014). Moreover, longitudinal studies demonstrate that greater hearing loss is related to greater gray matter volume loss (Lin et al., 2014; Xu et al., 2019) and lateral ventricle expansion (Eckert et al., 2019). These findings are consistent with the sensory deprivation hypothesis, such that less and/or degraded sensory input to the brain may lead to long-term deprivation effects on the auditory pathways causing structural decline as well as neurofunctional changes (Peelle and Wingfield, 2016). For example, Lin et al. (2014) showed that, in cognitively normal older adults, those greater hearing loss exhibited accelerated volume loss compared to those with normal hearing. Such accelerated volume loss was observed in several regions of the temporal lobe (i.e., right superior, middle, and inferior temporal gyri and the parahippocampus) that are important for auditory processing, semantic memory functioning, and cognitive processing. The associations between hearing loss and brain atrophy provide evidence for the sensory deprivation hypothesis because they suggest that long-term hearing loss is associated with specific types of structural loss in the brain. In older adults with dementia, hearing loss might even be directly interacting with dementia neuropathology in the medial temporal lobe – a hypothesis which has been put forward recently (Griffiths et al., 2020).

However, previous studies of the associations between hearing loss and brain atrophy have examined mainly healthy older adults with no clinically significant cognitive impairment. Studies of individuals who are particularly at risk for developing AD, as well as in those who have already been diagnosed with AD, could reveal whether hearing loss is independently associated with brain atrophy in those who typically have more vulnerable brains because of neuropathology-related atrophy. AD neuropathology typically causes accelerated gray and white matter declines which are core biomarkers of AD (for a review see Pini et al., 2016). Furthermore, individual differences in cognitive performance correlate with brain structural measures in those with subjective cognitive decline (SCD) and mild cognitive impairment (MCI). For example, lower memory performance in a face-name recall test, which is sensitive to impairment in early stages of AD, correlated with smaller right hippocampal volume in SCD and MCI (Caillaud et al., 2019). SCD refers to individuals who have subjective complaints about their cognitive capacities, but who perform within normal limits on behavioral neuropsychological tests (Jessen et al., 2010, 2014). MCI refers to individuals who show clinically significant impairment in one or more cognitive domains, while their functional abilities in everyday life are judged to be intact (Albert et al., 2011). There is longitudinal evidence that SCD is a risk factor for cognitive decline as well as for AD and that it occurs at the preclinical stage of AD and other dementias (Glodzik-Sobanska et al., 2007; Jessen et al., 2010; van Oijen et al., 2007; Reisberg et al., 2010). Similarly, MCI is a strong risk factor for AD and, in some individuals, describes an intermediate stage between normal cognitive aging or preclinical AD and AD. However, not all persons with MCI convert to AD, with reported rates between 20-40% (Albert et al., 2011; Roberts and Knopman, 2013).

The extent to which hearing loss is independently associated with (sub)cortical gray matter loss in those with varying degrees of cognitive impairment and neuropathology has yet to be examined. There is little evidence that pathological features of AD, such as amyloid plaques or neurofibrillary tangles, are observed in the cochlea (Sinha et al., 1993; Wang and Wu, 2015, but see Omata et al., 2016). Thus, an independent association between hearing loss and brain atrophy in those at risk for or with AD would be evidence for the sensory deprivation hypothesis in these individuals.

In the current study, we analyzed data from the first wave of data released from the COMPASS-ND (<u>Comp</u>rehensive <u>Ass</u>essment of <u>N</u>eurodegeneration and <u>D</u>ementia) study (Chertkow et al., 2019). The COMPASS-ND cohort includes participants with varying types and degrees of cognitive impairment (for more information about COMPASS-ND see: http://ccna-ccnv.ca/compass-nd-study/). We compared these diagnostic groups on measures of pure-tone hearing loss and speech-in-noise thresholds. We also examined the associations between the two auditory measures and expected them to be positively associated in each diagnostic group. Further, we examined the extent to which hearing loss is related to gray matter atrophy in the hippocampus as well as whole-brain analysis of cortical thickness in those with SCD, MCI, and AD. We hypothesized that higher (worse) pure-tone thresholds and/or higher (worse) speech-in-noise thresholds would be associated with lower cortical thickness in all three groups. Specifically, we expected pure-tone thresholds to be negatively correlated with cortical thickness in primary and secondary auditory areas, hippocampal volume, and possibly the prefrontal cortex (due to reallocation of resources). Furthermore, we hypothesized that speech-in-noise word recognition thresholds to be negatively associated with the cortical thickness of primary and secondary auditory areas, the prefrontal cortex, and the temporo-parietal areas involved in speech processing. Such associations would be evidence for the sensory deprivation hypothesis in those with or at high risk for developing dementia.

## 2. Material and Method

### 2.1. Participants

Participants in the present study were selected from those whose data were included in the first or second wave of the COMPASS-ND data released in November 2018 and May 2019, respectively. General COMPASS-ND inclusion criteria included: being between 50-90 years of age, having a study partner who sees the participant weekly and who can participate as required by the protocol, passing the safety requirements for the MRI scanning, and possessing sufficient proficiency in English or French (as judged by the examiner) to undertake self-report and neuropsychological testing. Exclusion criteria were as follows: presence of significant known chronic brain disease unrelated to AD, on-going alcohol or drug abuse which, in the opinion of the investigator, could interfere with the person’s ability to comply with the study procedures, severe cognitive impairment indicated by a score of < 13/30 on the Montréal Cognitive Assessment (MoCA; Nasreddine et al., 2005) or a symptomatic stroke within the previous year. Written informed consent was obtained from all participants. The COMPASS-ND study was approved by all relevant Research Ethics Boards.

In the present study, we analyzed the data from individuals who met the criteria for SCD (N=35), MCI (N=85), or AD (N=25). Of those 145 participants, 10 were excluded: four (3 MCI, 1 AD) did not have MRI data, one (MCI) did not have hearing data, and five (2 MCI, 3 AD) were considered to be outliers with measures that were +3 standard deviations (SD) above average in cortical volume. Thus, a final total of 135 participants (N=35 SCD; N=79 MCI; N=21 AD) were included. Table 1 provides an overview of the demographic and health variables for each group. We found significant group differences in age and sex, and the group difference in education approached significance (*p*=.1); thus, we included these variables as covariates in all analyses.

**Table 1:** Summary statistics on demographic, vision, and health variables for COMPASS-ND participants by diagnostic groups.

|  | SCD<br>(N=35) |  | MCI<br>(N=79) |  | AD<br>(N=21) |  |  |  |  |
| --- | --- | --- | --- | --- | --- | --- | --- | --- | --- |
|  | N |  | N |  | N |  | X<sup>2</sup> | p |  |
| Female | 20 (57 %) |  | 29 (37%) |  | 1 (5%) |  | 15.35 | **<.001** |  |
| Hearing aid users | 2 (6%) |  | 13 (17%) |  | 4 (19%) |  | 2.95 | .23 |  |
| Pocket talker | 1 (3%) |  | 3 (4%) |  | 2 (10%) |  | 1.56 | .46 |  |
| Right handedness | 32 (91%) |  | 76 (96%) |  | 19 (91%) |  | 7.30 | .12 |  |
| Smoking | 19 (54%) |  | 33 (45%) |  | 12 (57%) |  | 5.22 | .27 |  |
| Hypertension | 7 (20%) |  | 28 (36%) |  | 9 (43%) |  | 2.68 | .61 |  |
|  | M | SD | M | SD | M | SD | F | p | Post-hoc |
| Age, years | 69.45 | 6.18 | 73.47 | 6.57 | 75.87 | 7.24 | 7.24 | **.001** | SCD<MCI,AD |
| Education (years) | 16.79 | 3.19 | 15.52 | 3.07 | 15.31 | 2.96 | 2.36 | .10 | - |
| MoCA (score/30) | 27.23 | 1.94 | 24.06 | 3.06 | 19.38 | 3.04 | 51.21 | **<.001** | SCD>MCI>AD |
| Visual contrast sensitivity (CS)*<br>log units** | 1.70 | .15 | 1.67 | .15 | 1.48 | .17 | 12.62 | **<.001** | SCD,MCI>AD |
| Reading Acuity (logMAR units)*** | .28 | .27 | .30 | .29 | .20 | .21 | 1.26 | .29 | - |

SCD=subjective cognitive decline, MCI=mild cognitive impairment, and AD=Alzheimer's dementia groups.
\*most of the study participants had normal CS (SCI: 87.5%, MCI: 93%, AD: 50%), while few had moderately impaired CS (SCI: 12.5%, MCI: 7%, AD: 50%); however, these levels of impairment are unlikely to interfere with test administration of visual stimuli
\*\* < 1 log CS = severe impairment, 1-1.5 log CS = moderate impairment, > 1.5 log CS = normal for age 60+
\*\*\* logMAR = logarithm of the minimum angle of resolution; logMAR < .30 (equivalent of better than 20/40) = normal acuity. logMAR .30 to .50 (20/40 to 20/60) = moderate visual impairment

### 2.2. Criteria for SCD

One core criterion for SCD is a self-experienced persistent decline in cognitive capacities in comparison to previous normal status, with the decline being unrelated to an acute event. This criterion was operationalized by two questions 1) “Do you feel like your memory or thinking is becoming worse?” and, if so, 2) “Does this concern you?” Only individuals who answered the two questions with “yes” were assigned to the SCD group (following Jessen et al., 2014). Further inclusion criteria for SCD was normal age- and education-adjusted performance on standardized cognitive tests (Chertkow et al., 2019), including a) a score of ≥ 25/30 on the MoCA (Nasreddine et al., 2005), b) a word list recall score of > 5 on the Consortium to Establish a Registry for Alzheimer’s Disease (CERAD), c) performance above Alzheimer’s Disease Neuroimaging Initiative (ADNI) education-adjusted cut-offs on the delayed recall of the Logical Memory Subtest of the Wechsler Memory Scale-3^rd^ ed. (WMS-III; Tulsky et al., 2003), and d) no symptoms (i.e., a zero) on the Clinical Dementia Rating (CDR; Hughes et al., 1982).

### 2.3. Criteria for MCI

Participants who reported, or whose informants reported, a concern regarding a change in the participant’s cognition were included in the MCI group if they were determined to be able to follow daily life routines independently (Chertkow et al., 2019) and met at least one of the following four criteria representing impairment in one or more cognitive domains (Albert et al., 2011): a) WMS-III Logical Memory Delayed Recall score < ADNI education-adjusted cut-offs, b) CERAD word list recall < 6, c) MoCA score 13-24/30, d) assigned a CDR of ≤ 0.5.

### 2.4. Criteria for AD

Participants who were diagnosed with AD were selected based on the following three criteria (following McKhann et al., 2011): a) a gradual progressive change in memory and/or other cognitive functions over more than six months based on the participant’s and/or informant’s report; b) objective evidence of a significant decline in at least two domains of cognition (i.e., episodic memory, reasoning, problem solving, visuospatial abilities, language, personality/behavior) as defined by fulfilling at least two of the following criteria: Logical Memory II score below ADNI cutoffs, CERAD word list recall < 7, MoCA score 13-24/30 (with at least one point lost in a non-memory task), or a positive response to the question: “Has the participant had any changes in personality or behavior?”; c) the presence of impairment in functional abilities operationalized by a positive response to the statement, “The cognitive deficits interfere with independence in everyday activities such as paying bills or managing medications” (Chertkow et al., 2019).

### 2.5. Hearing loss

#### 2.5.1. Pure-tone audiometry

Pure-tone audiometry was conducted using an abbreviated screening protocol using a GSI 18 audiometer in a quiet clinical examination room. The screening protocol assessed if participants were able to detect at least one of two presentations of a pure tone presented at each of 3 pre-selected frequencies at fixed dB HL levels. Each ear was tested separately. First, there were two trials where a 2-kHz pure tone was presented at 40 dB HL. If the participant successfully detected at least one presentation at 40 dB HL, then two trials at 25 dB HL were presented at 2 kHz, 1 kHz, and 4 kHz. Participants who failed to hear a 2-kHz pure tone at 40 dB HL were provided with a Pocket Talker assistive listening device throughout the neuropsychological and clinical assessment if they did not have their own hearing aid.

For the present analyses, participants were assigned to one of six hearing loss categories based on their ability to detect at least one of the 2-kHz pure tones at 40 dB HL or 25 dB HL as described in the left-hand column of Table 2^2^. Using the audiometric thresholds of older adults from two large datasets, the six categories defined based on the COMPASS-ND pure-tone screening protocol were validated by examining their correspondence to commonly used categories of hearing loss that are typically determined by four-frequency pure-tone averages (see Table 2). In order to validate the six categories of hearing loss defined based on the pure-tone screening protocol used in COMPASS-ND based on the detection of a 2-kHz pure tone at 25 or 40 dB HL, we applied the same classification method to two large databases for older adults whose audiometric thresholds had been measured. The first dataset was a community-based sample of 27,444 healthy Canadians who participated in the baseline comprehensive cohort of the Canadian Longitudinal Study on Aging (CLSA) (Raina et al., 2009), for whom air-conduction pure-tone audiometric thresholds were measured at octave frequencies from .25 to 8 kHz in each ear (CLSA data release 3.2; see https://www.clsa-elcv.ca/doc/529 for CLSA audiometry protocol). We categorized these participants according to which screening category their PTA results would place them in and then, within each category, calculated their mean pure-tone average (PTA). Table 2 shows the 1, 2, 3, and 4 kHz PTA for the CLSA comprehensive cohort based on the COMPASS-ND classification scheme described above. There was a monotonic worsening in PTA from Category 1 (better hearing) to Category 6 (worse hearing), confirming that the COMPASS-ND method properly distinguishes people according to their hearing abilities.

**Table 2:**
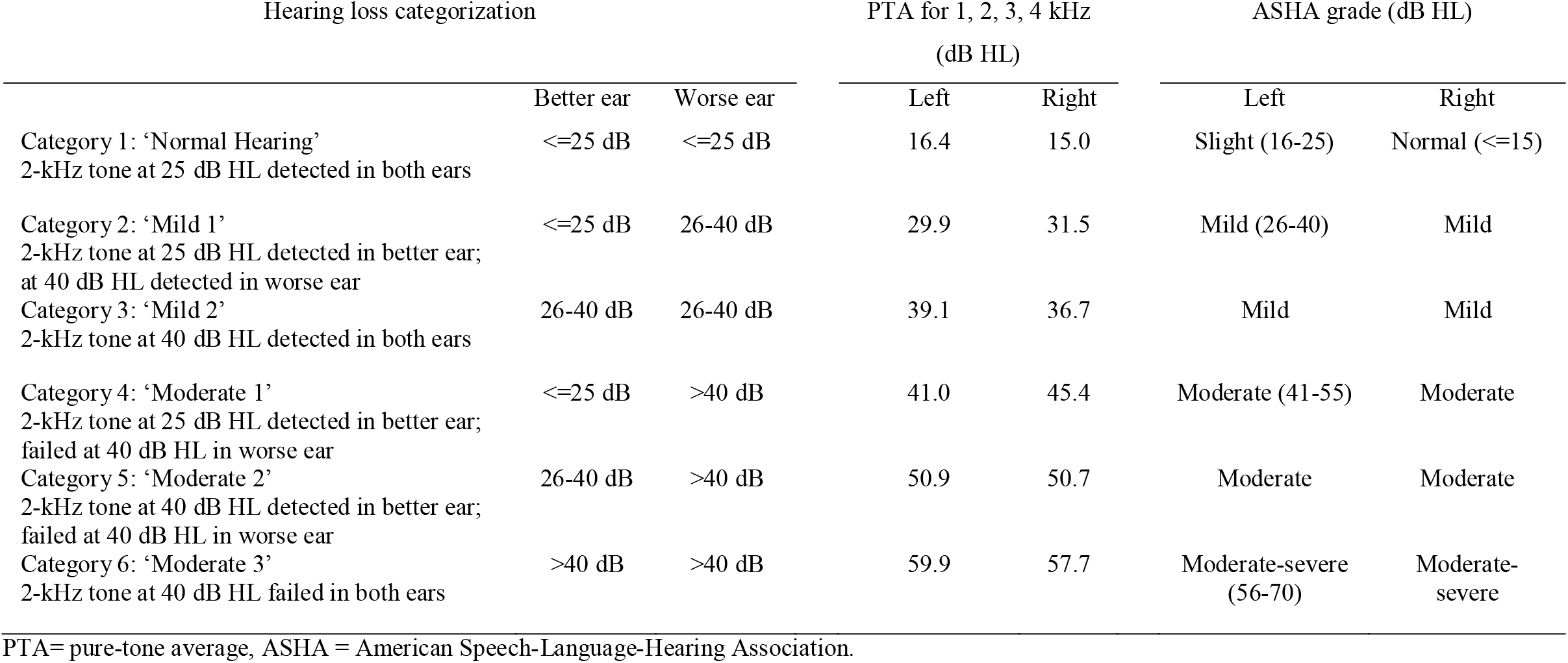
Validation of the 6-level hearing classification system used in the current study. Table 2 demonstrates the pure-tone averages (1, 2, 3, and 4 kHz) of participants in the Canadian Longitudinal Study on Aging when the classification system was applied to them.

| Hearing loss categorization |  |  | PTA for 1, 2, 3, 4 kHz<br>(dB HL) |  | ASHA grade (dB HL) |  |
| --- | --- | --- | --- | --- | --- | --- |
|  | Better ear | Worse ear | Left | Right | Left | Right |
| Category 1: ‘Normal Hearing’<br>2-kHz tone at 25 dB HL detected in both ears | <=25 dB | <=25 dB | 16.4 | 15.0 | Slight (16-25) | Normal (<=15) |
| Category 2: ‘Mild 1’<br>2-kHz tone at 25 dB HL detected in better ear;<br>at 40 dB HL detected in worse ear | <=25 dB | 26-40 dB | 29.9 | 31.5 | Mild (26-40) | Mild |
| Category 3: ‘Mild 2’<br>2-kHz tone at 40 dB HL detected in both ears | 26-40 dB | 26-40 dB | 39.1 | 36.7 | Mild | Mild |
| Category 4: ‘Moderate 1’<br>2-kHz tone at 25 dB HL detected in better ear;<br>failed at 40 dB HL in worse ear | <=25 dB | >40 dB | 41.0 | 45.4 | Moderate (41-55) | Moderate |
| Category 5: ‘Moderate 2’<br>2-kHz tone at 40 dB HL detected in better ear;<br>failed at 40 dB HL in worse ear | 26-40 dB | >40 dB | 50.9 | 50.7 | Moderate | Moderate |
| Category 6: ‘Moderate 3’<br>2-kHz tone at 40 dB HL failed in both ears | >40 dB | >40 dB | 59.9 | 57.7 | Moderate-severe<br>(56-70) | Moderate-severe |

PTA= pure-tone average, ASHA = American Speech-Language-Hearing Association.

The second dataset was obtained from the another project (N=242, mean age = 70.67 (SD = 5.94), mean years of education = 15.25 (SD = 2.31), 69.4% women), to investigate stigma related to aging and hearing loss in Canada, to correlate our hearing loss categorization variable with the PTA of full audiograms (1000, 2000, 4000 Hz) of the better ear (*r*=.84, *p*<.001) and the worse ear (*r*=.85, *p*<.001). Overall, these two additional datasets show that our hearing loss categorization based on the more restricted COMPASS-ND hearing screening protocol was valid.

For the statistical analyses we treated the six hearing loss categories as scaled data, because Categories 1-6 reflect the degree of hearing loss.

#### 2.5.2. Canadian Digit Triplet Test (CDTT)

The Canadian Digit Triplet Test (CDTT) was used to measure participants’ speech reception threshold (SRT) corresponding to the signal-to-noise ratio at which triplets of digits are recognized correctly 50% of the time. Participants were instructed to listen to three digits presented in speech-shaped background noise and to repeat them. The CDTT application was run on a Dell XPS laptop using a USB audio card (Creative Sound Blaster X-Fi Go! Pro). The CDTT uses an adaptive 1-up-1-down procedure; the speech level increases after a correct response (all three digits repeated correctly) and decreases after an incorrect response (Ellaham et al., 2016; Giguère et al., 2020). For each participant, the standard deviation (SD) of the responses and the number of reversals were used to identify erratic runs. SRTs from all participants were considered be valid with the exception of 2 participants (1 AD, 1 MCI) who had SDs higher than +3 above the mean SD for each diagnostic group and whose results were excluded from analyses. Furthermore, the SRT of 1 additional participant (SCD), who was categorized as having “Mild 1” pure-tone hearing loss based on audiometric screening results, was excluded because this individual’s SRT was higher than +4 dB SNR which is atypical for persons with mild hearing loss in this sample (this participant)^3^.

### 2.6. MRI data acquisition and analyses

T1-weighted images were obtained from each participant using 3T scanners following the Canadian Dementia Imaging Protocol (CDIP) (Duchesne et al., 2019). The CDIP is a validated, harmonized protocol for multi-site MRI data acquisition to study neurodegeneration and is available for scanners manufactured by major vendors (GE, Phillips, and Siemens). The parameters for the acquisition of the 3D T1-weighted images can be found here https://www.cdip-pcid.ca/. The CDIP established parameters for each scanner type and version allowing for the images to be as comparable as possible across scanners.

Hippocampal volumes were extracted from the T1-weighted images which were submitted to the ANIMAL (automatic non-linear image matching and anatomical labeling) segmentation method which is an atlas-based segmentation method that uses non-linear registration to a pre-labeled template (Collins and Pruessner, 2010; Collins et al., 1995). Furthermore, we performed a FreeSurfer analysis (version 4.2, http://freesurfer.net/) on the T1-weighted images of the same participants to extract cortical thickness of cortical regions. The FreeSurfer pipeline performs surface-based morphometry (SBM) which involves several processing steps that have been described in detail in previous publications (Dale et al., 1999; Fischl and Dale, 2000; Fischl et al., 1999, 2002, 2004). The fully automated pipeline was run on CBRAIN (Sherif et al., 2014), a web-based software for computing intensive analyses that generates individual cortical surface models with high spatial precision. Data from five MCI participants were excluded due to errors during preprocessing. All other brain scans were manually checked for the segmentation precision, resulting in 3 additional MCI participants being removed from the analysis due to severe segmentation errors.

Cortical thickness was measured by the minimal distance between the gray-white matter border and the pial surface. FreeSurfer-based cortical thickness values have previously been validated against manual measurements of cortical thickness as well as histological analysis (Cardinale et al., 2014) and have been shown to be reliable in healthy older adults (Liem et al., 2015). Whole-brain statistical analyses were calculated using the FreeSurfer built-in general linear model (GLM) toolbox to identify thickness of brain regions associations with hearing loss category (GLM 1) and CDTT (GLM 2). In order to compute the GLMs, each participant’s segmented brain was morphed to an average spherical surface and smoothed using a FWHM kernel of 15 mm. A significance threshold of *p*=.001 was applied as was done in previous studies on older adults with hearing-related disorders such as tinnitus (Meyer et al., 2016; Vanneste et al., 2015).

### 2.7. Statistical analyses and covariates

We used age, sex, and education as covariates in all statistical analyses. When exploring the relationship between CDTT SRT and cortical thickness, we controlled for pure-tone hearing loss category. IBM SPSS Statistics 24 was used for statistical analyses.

Three families of statistical analyses were conducted as follows: First, two one-way ANOVAs (Section 3.1.) were conducted with diagnostic group as a between-subjects variable to determine if the groups differed in their degree of audiometric hearing loss using hearing loss category and subsequently CDTT SRT as the dependent variables. Second, the association between the two different measures of hearing loss (i.e., audiometric hearing loss category and CDTT SRT) was explored within each diagnostic group using parametric partial correlations (Section 3.2.). Third, associations between hearing loss and hippocampal volumes were computed using parametric partial correlations. Associations between hearing loss and cortical data were computed as described above (Section 3.3.). Unless otherwise indicated, an alpha level of α = 0.05 was accepted and effect sizes were indicated by partial eta squares (η _p_^2^).

## 3. Results

### 3.1. Diagnostic group comparisons in hearing

Figure 1 shows the audiometric hearing loss categories and CDTT SRT measures for each of the three diagnostic groups. The diagnostic groups did not differ significantly on the audiometric hearing loss category measure (*F*(2,126)=.33, *p*=.72, η_*p*_^*2*^=.005; Figure 1, left panel). Most participants were categorized as having normal audiometric thresholds or mild hearing loss, while a notable minority had moderate hearing loss or greater. There was a trend towards diagnostic groups differing on CDTT SRT (*F*(2,109)=2.73, *p*=.07, η_*p*_^*2*^=.05), see Figure 1 right-hand side. Post-hoc t-tests showed a trend towards higher (poorer) SRTs in the AD compared to the SCD group, with this difference between groups approaching but not reaching significance (*p*=.08).

**Figure 1:**
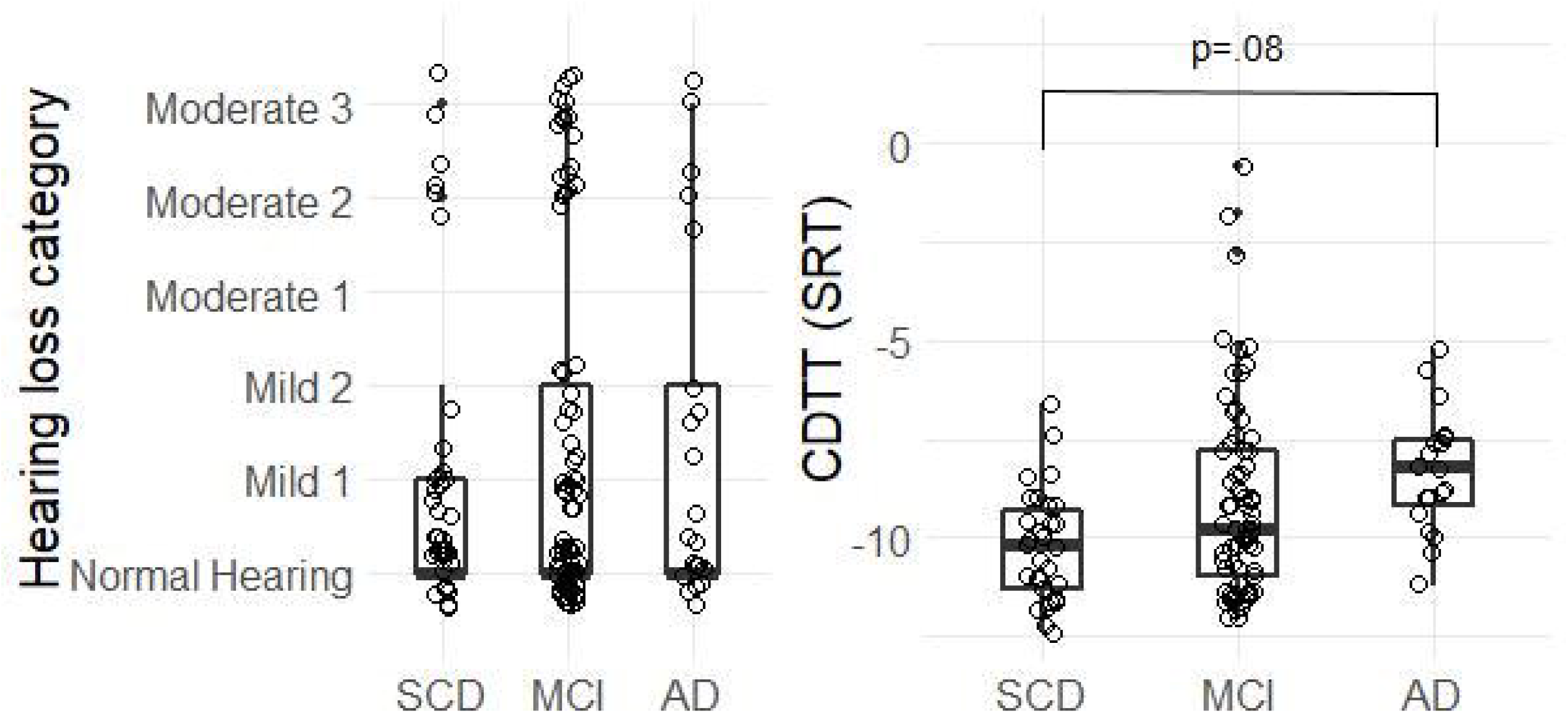
Figure 1 depicts the pure-tone hearing loss categories and the CDTT SRTs for participants in the three diagnostic groups (SCD: subjective cognitive decline; MCI: mild cognitive impairment; AD: Alzheimer’s dementia). There was no significant difference in pure-tone hearing loss category between the three cognitive diagnostic groups, but the ANOVA indicated differences in CDTT SRTs between groups which were higher (worse performance) in the AD compared to the SCD group (controlling for age, sex, education, and HL category). The boxplot lines represent the group median (thick line) and the 25^th^ and 75^th^ percentiles (outer lines). Dots represent results for individual participants.

### 3.2. Associations between audiometric hearing loss category and CDTT SRT

Partial correlations between hearing loss category and CDTT thresholds (see Figure 2) revealed significant positive associations (based on an adjusted *p*-value divided by three for each correlation computed in the three diagnostic groups: .05/3=.017) in the SCD group (*r*(24)=.64, *p*<.001, R^2^=.41) and the MCI group (*r*(63)=.70, *p*<.001, R^2^=.49), and associations approaching significance in the AD group (*r*(14)=.43, *p*=.07, R^2^=.18), suggesting that the more pure-tone hearing loss the participants exhibited, the greater (worse) their SRTs were on the CDTT. Furthermore, even though the association was only trending in the AD group, the Fisher r-to-z transformation of the correlation coefficients suggested that the strengths of the associations were not different between the diagnostic groups (SCD vs. MCI *p*=.56, SCD vs. AD *p*=.36, MCI vs. AD *p*=.58).

**Figure 2:**
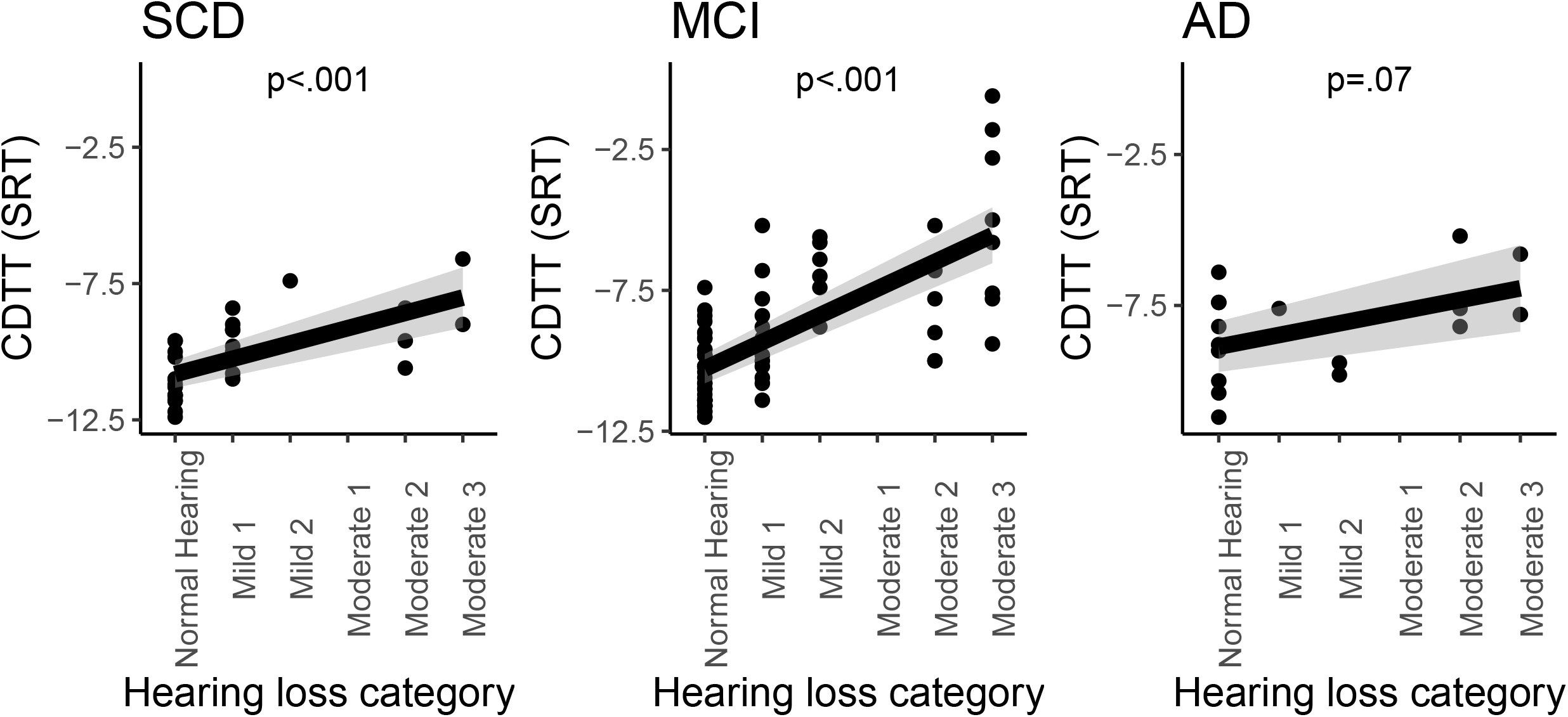
Figure 2 shows the associations between pure-tone hearing loss category and the CDTT SRTs for the three diagnostic groups (subjective cognitive decline (SCD), mild cognitive impairment (MCI), and Alzheimer’s dementia (AD)). Significant positive associations were found in the SCD group (R^2^=.41) and the MCI group (R^2^=.49), and associations approached significance (*p* < .1) in the AD group (R^2^=.18). Grey areas indicate 95% confidence intervals.

### 3.3. Associations between hearing measures and brain structural measures

#### 3.3.1. Hearing loss and hippocampal volume

Table 3 and Figure 3 show the associations between the two hearing loss measures and hippocampal volume. SCD participants who had greater pure-tone hearing loss had lower gray matter volume in the right hippocampus (R^2^=.17). No significant correlations were found in the MCI and AD groups, suggesting that for those with stronger cognitive impairment, the association between pure-tone hearing loss and brain structure was not reliable. In order to statistically assess the observation that the correlation coefficients between pure-tone hearing loss and right hippocampal volume are different for diagnostic groups (i.e., greater pure-tone hearing loss is associated with lower right hippocampal volume in only those without objective evidence of cognitive impairment), we calculated a Fisher r-to-z transformation of the correlation coefficients. The findings supported this interpretation with SCD vs. MCI *p*=.04., SCD vs. AD *p*=.45, MCI vs. AD *p*=.10.

**Table 3:** Parametric partial correlations (controlling for age, sex, and education) between pure-tone hearing loss category or CDTT SRTs (controlled also for hearing loss category) and hippocampal volume are shown separately for each cognitive diagnostic group.

|  |  | SCD |  | MCI |  | AD |  |
| --- | --- | --- | --- | --- | --- | --- | --- |
|  |  | Left HC | Right HC | Left HC | Right HC | Left HC | Right HC |
| Hearing loss category | *r* | -.14 | **-.41** | .09 | -.07 | -.30 | -.38 |
|  | *p* | .46 | .03 | .47 | .56 | .26 | .14 |
|  | *R*<sup>2</sup> | .02 | .17 | .01 | .00 | .09 | .14 |
| CDTT | *r* | .02 | -.02 | .08 | .00 | .01 | -.52 |
|  | *p* | .93 | .92 | .53 | .98 | .97 | .07 |
|  | *R*<sup>2</sup> | .00 | .00 | .01 | .00 | .00 | .27 |

Bold indicates $p < .05$ . HC=hippocampus. SCD=subjective cognitive decline, MCI=mild cognitive impairment, AD=Alzheimer's dementia.

**Figure 3:**
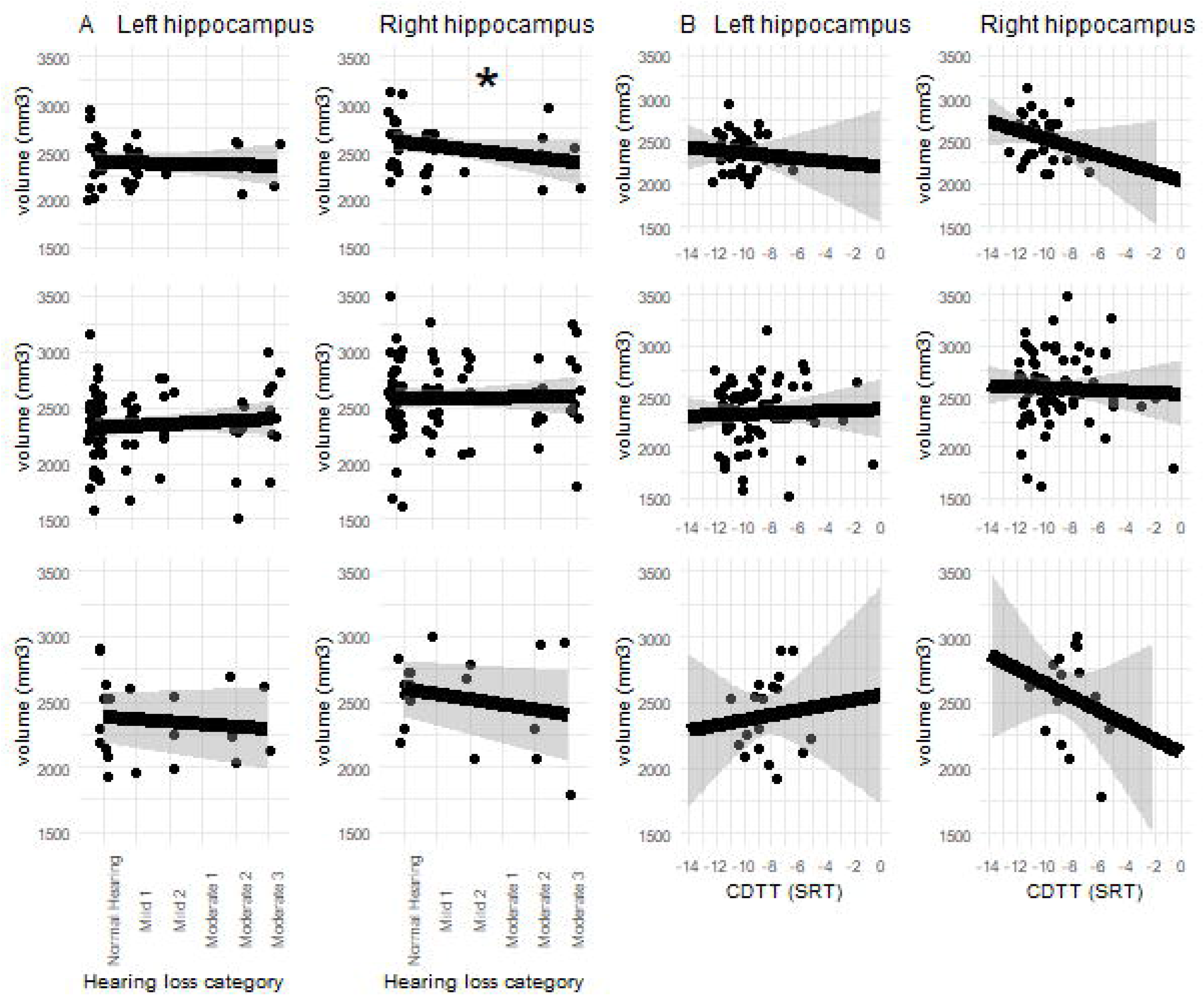
Figure 3 shows the correlations between pure-tone hearing loss category (A) or the CDTT SRTs (B) and hippocampal volume (mm^3^). In the subjective cognitive decline (SCD) group (top), there was a significant association, with greater hearing loss associated with lower right hippocampal volume, marked with colored square (\**p* < .05). No significant associations between pure-tone hearing loss category or the CDTT SRTs and the hippocampal volume were found in MCI and AD. Grey areas indicate 95% confidence intervals. Also shown are results for the MCI (mild cognitive impairment) group (middle panel) and the AD (Alzheimer’s dementia) group (lower panel).

#### 3.3.2. Hearing loss and cortical thickness

The whole-brain GLM model showed that there were two significant positive associations between hearing loss category and cortical thickness, one in the left superior temporal gyrus and the other in the right pars opercularis (*p*<.001, controlling for multiple comparisons) as shown in Figure 4 and Table 4. Both significant associations were observed in the SCD group, with those who had greater (worse) pure-tone hearing loss having greater cortical thickness in those two brain regions. No significant associations were found in the MCI or AD groups once we controlled for multiple comparisons.

**Table 4:** Table 4 shows the signifcant assocations between pure-tone hearing loss category or the CDTT SRTs and cortical thickness for the three diagnostic groups: subjective cognitive decline (SCD), mild cognitive impairment (MCI), and Alzheimer’s dementia (AD). The log10(p) at each vertex was set to 3 (values > 3 correspond to *p* < .001).

|  |  |  | Correlation | Max | x | MNI<br>y | z |
| --- | --- | --- | --- | --- | --- | --- | --- |
| Hearing loss category |  |  |  |  |  |  |  |
| SCD | LH | Superior temporal gyrus | positive | 3.06 | -39.88 | -20.29 | -20.40 |
|  | RH | Pars opercularis | positive | 3.75 | 21.98 | 39.72 | -10.32 |
| MCI | - | - | - | - | - | - | - |
| AD | - | - | - | - | - | - | - |
| CDTT |  |  |  |  |  |  |  |
| SCD | LH | Posterior cingulate cortex | positive | 3.98 | 37.97 | -8.41 | 13.51 |
|  | RH | Superior temporal gyrus | positive | 3.08 | 37.77 | -28.2 | -25.35 |
| MCI | LH | Supramarginal gyrus | positive | 3.10 | -32.09 | -47.51 | 24.74 |
|  | RH | Superior frontal gyrus | positive | 3.45 | -31.23 | 79.16 | 9.91 |
|  |  | Lateral orbitofrontal gyrus | positive | 3.52 | 0.92 | 72.74 | -45.23 |
|  |  | Anterior cingulate gyrus | positive | 3.25 | -33.03 | 61.90 | 14.18 |
| AD | LH | Superior temporal gyrus | negative | -3.30 | -39.88 | -20.29 | -20.40 |
|  | RH | Superior parietal gyrus | negative | -3.50 | -18.38 | -95.97 | 9.31 |
|  |  | Superior temporal sulcus | negative | -3.34 | 37.77 | -28.2 | -25.35 |
|  |  | Lateral occipital gyrus | negative | -3.32 | 10.43 | -92.77 | -20.07 |
|  |  | Inferior temporal gyrus | negative | -3.05 | 18.67 | -11.54 | -64.7 |

LH: Left hemisphere, RH: Right hemisphere, Max: log10(p) at peak, Montreal Neurological Institute (MNI) coordinates.

**Figure 4:**
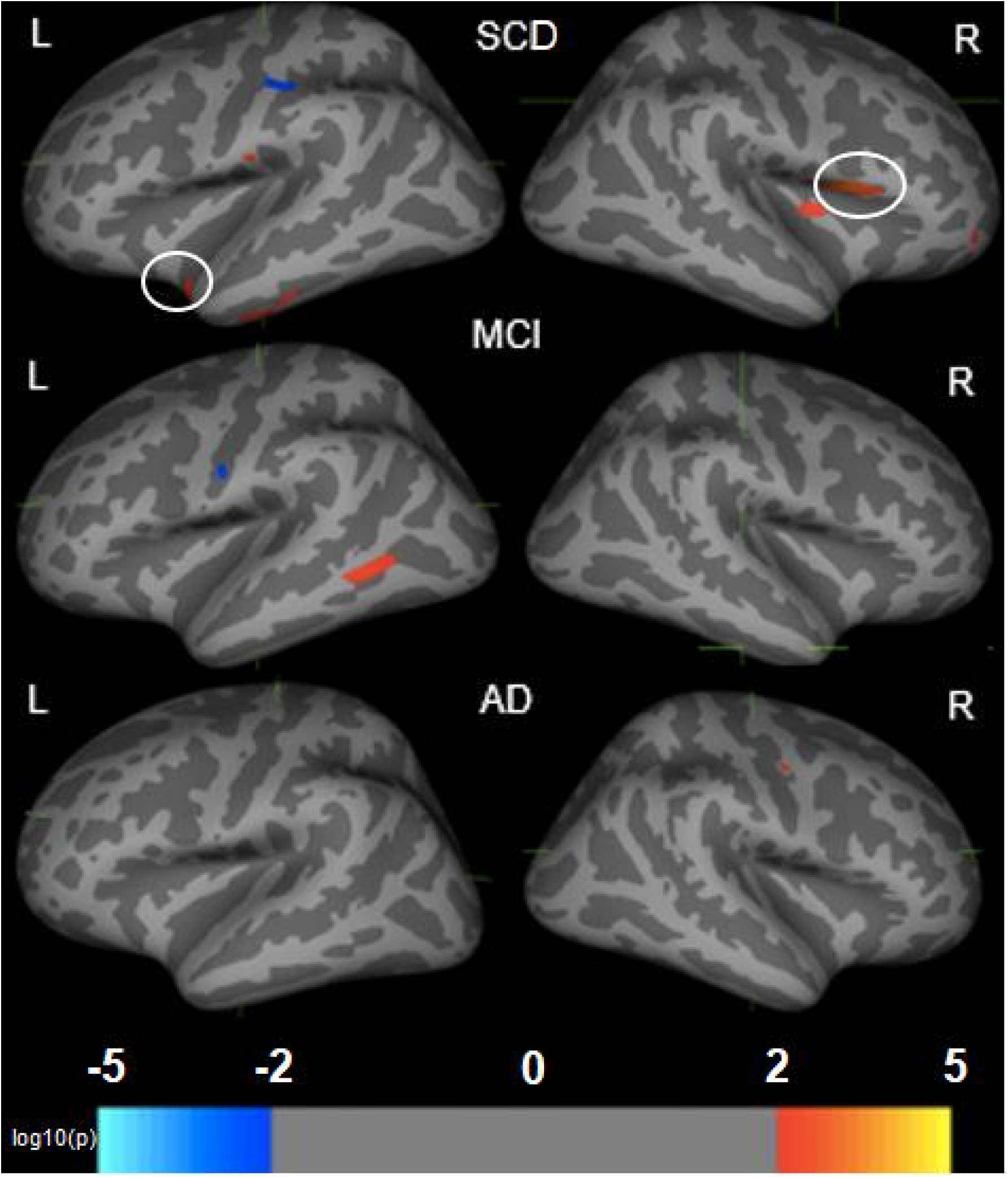
Figure 4 depicts the signifcant assocations between pure-tone hearing loss category and cortical thickness (CT) in the three diagnotic groups (subjective cognitive decline (SCD); mild cognitive impairment (MCI); Alzheimer’s dementia (AD)). Red = positive association. Blue = negative association. For graphical purposes, the log10(p) at each vertex was set to 2 (values > 2 correspond to *p* < .01), while the *p*-value was lowered to *p* < .001 to control for multiple comparisons in Table 5 where the significant brain clusters are listed. Significant (*p* < .001) positive correlations (i.e., greater pure-tone hearing loss and greater CT) were found in the SCD group for the left superior temporal gyrus and the right pars opercularis (indicated with circles).

The whole-brain GLM model using CDTT SRT as a predictor resulted in several significant clusters across the brain, as shown in Figure 5 and Table 4. In the SCD and MCI groups, a few positive correlations were found. In contrast, in the AD group, there were negative correlations in temporal, parietal, and occipital brain regions, indicating that those who have greater (worse) CDTT SRTs have less cortical thickness in a number of brain regions across the cortex.

**Figure 5:**
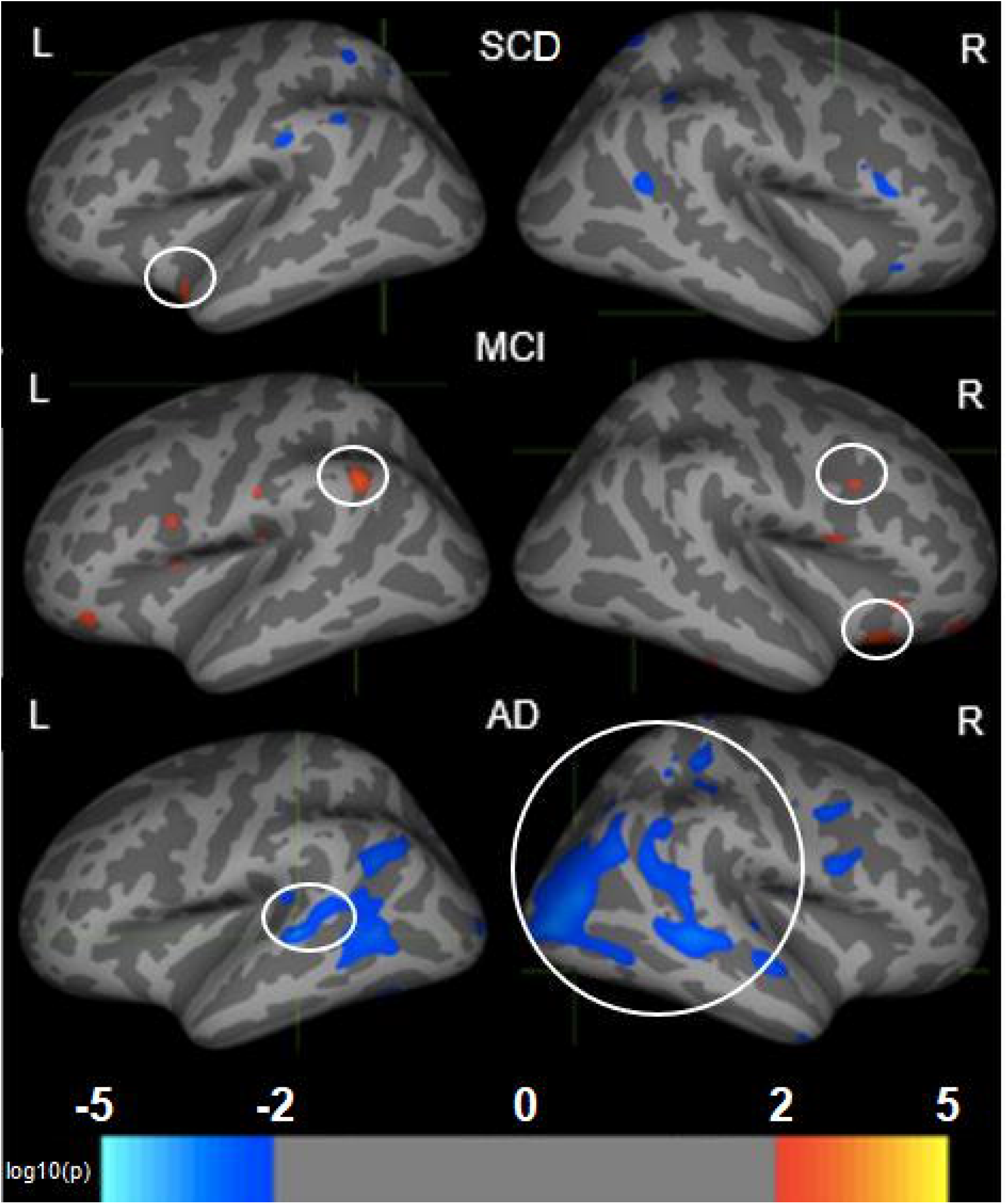
Figure 5 shows the signifcant assocations between CDTT SRTs and cortical thickness (CT) in the three diagnotic groups (subjective cognitive decline (SCD); mild cognitive impairment (MCI); Alzheimer’s dementia (AD)). Red = positive association. Blue = negative association. The log10(p) at each vertex was set to 2 for the figure (values > 2 correspond to *p* < .01), while the *p*-value was lowered to *p* < .001 to control for multiple comparisons in Table 5 where the significant brain clusters are listed. Significant associations at *p* <.001 were found as indicated with circles.

## 4. Discussion

The findings of the present study indicate that there are significant associations between hearing loss and brain anatomical measures. Importantly, these associations differed by diagnostic group and whether hearing loss is measured in terms of pure-tone detection or the supra-threshold CDTT speech-in-noise SRT measure. In persons with cognitive complaints (i.e., SCD), pure-tone hearing loss category was associated with reduced right hippocampal volume (see Figure 3) and increased cortical thickness in the left superior temporal gyrus and the right pars opercularis (see Figure 4). In individuals with MCI, CDTT speech-in-noise SRTs were associated with increased cortical thickness in the right supramarginal gyrus, the superior frontal gyrus, the orbitofrontal gyrus and the anterior cingulum (see Figure 5). For participants with diagnosed AD, the CDTT SRTs were associated with wide-spread loss in cortical thickness (see Figure 5). The significance and implications of these findings are discussed in detail in the next sections.

### 4.1. Associations between hearing category based on pure-tone detection and brain structure differ across diagnostic groups

Our diagnostic groups did not differ in hearing loss category based on the detection of pure tones when controlling for age, sex, and education. Importantly, this suggests that associations between pure-tone hearing loss categories and specific brain structure are not confounded by group differences in the degree of hearing loss. Nevertheless, the groups differed in the associations between pure-tone hearing loss category and neuroanatomical measures.

Despite the fact that the groups did not differ in their degree of hearing loss, we found that more severe pure-tone hearing loss was associated with more cortical atrophy in the right hippocampus, but only in the SCD group. Notably, based on the amount of variance explained, our results indicate that SCD participants with moderate hearing loss (those in hearing loss categories 5 or 6) have 4% lower volume in the right hippocampus than SCD participants with normal hearing or mild hearing loss (those in hearing loss categories 1, 2, or 3), even after controlling for age, sex, and education. In other words, the right hippocampus of these individuals with moderate-to-severe hearing loss “looks” approximately 8 years older given that the normal shrinkage is approximately 0.5% annually for people above 60 years of age (Fjell et al., 2009). Previous cross-sectional research has found similar associations between pure-tone hearing loss and hippocampal volume in healthy older adults (Uchida et al., 2018). There have been similar findings in longitudinal studies showing accelerated decline in the hippocampus (Xu et al., 2019) and in the right parahippocampus (Lin et al., 2014) across time in healthy older adults with pure-tone hearing loss. Our results therefore extend previous findings by showing that the association between hearing loss and hippocampal volume is also evident in older adults who subjectively believe that their cognition has declined and who are at risk of developing Alzheimer’s disease, even though their performance on neuropsychological tests is considered to be normal. Future research should investigate a possible link between hearing loss, cognition, and hippocampal volume, given that cognitively healthy older adults with greater pure-tone hearing loss perform worse on memory tasks such as a) in (delayed) word recall (Colsher and Wallace, 1990; Deal et al., 2015; Ray et al., 2018) and b) in the free and cued selective reminding test (FCSRT) (Lin et al., 2011b)). Performance in such tasks is mediated by the hippocampus (e.g. word recall (Fernández et al., 1999) and FCSRT (Slachevsky et al., 2018)), while there are also sex differences to be considered (Al-Yawer et al., submitted).

Interestingly, the associations between pure-tone hearing loss category and cortical thickness in the SCD group were mostly positive, meaning that those with greater pure-tone hearing loss had more cortical thickness in the left superior temporal gyrus (STG) and the right pars opercularis. Previous studies that have found positive relationships between degree of pure-tone hearing loss and gray matter volume have interpreted the results within a compensation framework (Alfandari et al., 2018). The left STG, belonging to the auditory association cortex, is involved in the spectro-temporal analysis of speech (Hickok and Poeppel, 2007) and multisensory integration of auditory and visual speech cues (Callan et al., 2001; Möttönen et al., 2002). Moreover, the structural integrity of the left STG is a predictor of auditory working memory (Leff et al., 2009). Therefore, it is possible that those who have greater pure-tone hearing loss rely more on multisensory cues (e.g., visual speech cues) and on working memory during speech understanding such that they show alterations in the structure of the STG as a function of pure-tone hearing loss. Similarly, the right pars opercularis is involved in speech processing (Vigneau et al., 2011), particularly in speech perception tasks involving vowel tone pitch discrimination (Joanisse and Gati, 2003), syllable discrimination (Poeppel et al., 2004), and sentence pitch and linguistic prosody processing (Meyer et al., 2002). Thus, it is possible that those with greater pure-tone hearing loss might rely more on prosodic speech cues (Giroud et al., 2018, 2019), leading to structural plasticity in the right pars opercularis.

In contrast to the significant associations between pure-tone hearing loss category and hippocampal volume as well as cortical thickness in SCD, we did not find any significant associations between pure-tone hearing loss category and hippocampal volume or cortical thickness in MCI or AD. The results suggest that the associations between pure-tone hearing loss and gray matter loss are undetectable or not significant in groups with greater cognitive impairment (i.e., MCI and particularly AD), even though they typically have strong reductions in gray matter as compared to the SCD group. This is consistent with the findings of Xu et al. (2019) who reported more rapid decline in the hippocampus in those with greater pure-tone hearing loss in the preclinical stage (i.e., when AD is clinically asymptomatic but biomarkers suggest the presence of amyloid pathology) and in MCI, but not in individuals already diagnosed with dementia. Nevertheless, it remains an open question as to whether persons with either MCI or AD who have pure-tone hearing loss show greater cognitive decline than those without pure-tone hearing loss.

In sum, our data did not reveal any differences in pure-tone hearing loss between diagnostic groups. In contrast, one previous study reported that there was a higher prevalence of hearing loss (> 35 dB HL based on PTA of .5, 1, 2, and 4 kHz in the better ear) in people with MCI compared to healthy older adults and in people with AD compared to those with MCI and healthy older adults (Quaranta et al., 2014); however, most studies do not have such data because they examined the association between pure-tone hearing loss and cognitive decline in healthy older adults or in a group who were initially healthy and at risk of developing dementia (Deal et al., 2015, 2016, 2019; de la Fuente et al., 2019; Okely et al., 2019; Osler et al., 2019).

Our cross-sectional analysis further indicates that moderate pure-tone hearing loss in those who are otherwise cognitively healthy, but who have subjective cognitive complaints, is associated with lower volume in the right hippocampus, a biomarker of dementia. We note that we did not find lower hippocampal volume as a function of pure-tone hearing loss in the MCI or AD group in our sample. Furthermore, those with greater pure-tone hearing loss in the SCD group also exhibited more cortical thickness in the left STG and the right pars opercularis. This finding suggests potentially compensatory structural plasticity insofar as those with greater pure-tone hearing loss may rely more on multisensory and prosodic speech cues as well as the phonological loop component in their working memory during speech understanding compared to those with less severe pure-tone hearing loss. The fact that these hearing-brain structure associations were found only in individuals with SCD, but not MCI or AD, suggests that dementia-related neuropathology, which is reflected in brain atrophy among other biomarkers, may be overshadowing the potential effects of pure-tone hearing loss on brain structure.

### 4.2. Associations between hearing loss and brain structure depend on hearing loss measurement

In addition to the association between measures of brain structure and hearing loss category based on the detection of pure tones, we also use the CDTT measure of speech-in-noise SRT as a predictor of cortical structure. First, independent of hearing loss category, there was a trend for CDTT SRTs to be lower (better) for the SCD group compared to the AD group. This pattern of results suggests that those with AD have slightly (although not significantly) more difficulty recognizing even very simple speech stimuli (digit triplets) in noise. This is consistent with findings of central auditory processing dysfunctions in individuals with AD and partially also in those with MCI on an auditory processing measure using dichotic presentation of digits (Idrizbegovic et al., 2011).

We also investigated the associations between CDTT SRTs and cortical thickness in all three diagnostic groups. For the AD group only, we found negative correlations between CDTT SRTs and brain structure across the whole brain. Specifically, those with greater (poorer) CDTT SRTs had lower cortical thickness in bilateral superior temporal gyri, right inferior temporal gyrus, right lateral occipital gyrus, and the right superior parietal gyrus. Previous research has found similar associations between measures of speech-in-noise perception and macro-anatomical measures of the cortex in healthy older adults, which were more focal in nature (Giroud et al., 2018, 2020; Rudner et al., 2019). Cortical thickness changes as a function of experience, training, pathology, and lifespan and are therefore subject to plasticity (Engvig et al., 2010; Fjell et al., 2009). It is possible that greater (poorer) CDTT SRTs in the AD group correspond to a lower number, packing density, and size of cells within neural columns (Rakic, 1995) in multiple brain regions. These results may support the sensory deprivation hypothesis insofar as continued impoverished auditory input may have wide-spread effects across the cortex that manifest as gray matter atrophy. Thus, increasing difficulties understanding speech-in-noise perception could lead to a structural decline in those brain regions, as suggested by a recent cross-sectional study in healthy older adults in which lower performance on a speech-in-noise task was related to lower gray matter volume in brain regions associated with hearing and cognition (Rudner et al., 2019).

An alternative, and equally possible explanation, derives from the fact that a large cortical-subcortical brain network supporting auditory-cognitive functioning is involved in speech-in-noise processing, ranging from the auditory brainstem (Anderson et al., 2011, 2013) up to higher areas such as the right inferior frontal gyrus and the right insula (Bidelman and Howell, 2016), including parietal brain regions such as the precuneus (Wong et al., 2009). These brain regions involved in speech-in-noise processing are similar to the brain structures we found were correlated with CDTT SRTs in the present study. Thus, it also is possible that adequate performance on the CDTT depends on intact brain structures involved in perceiving speech in noise. In other words, it is possible that individuals with reduced gray matter volume and cortical thickness due to neurodegeneration (i.e., those with AD who have more atrophy) in those critical brain regions perform worse on the speech-in-noise task due to less available neural resources. However, longitudinal research and functional imaging is needed to clarify the direction of dependency.

Similar to the study of Giroud et al. (2018) with healthy older adults, we also found associations between performance on a speech-in-noise measure and brain structures in visual brain areas (i.e., volume in the right occipital lobe and cortical thickness in the right lateral occipital gyrus). Visual system activation occurs during hearing tasks (Giraud and Truy, 2002), especially in individuals with severe hearing loss such as those using cochlear implants (Giraud et al., 2001). It is possible that individuals with greater hearing loss rely more on visual speech cues and therefore co-activate visual areas during speech understanding to a greater extent than those with normal hearing, potentially even when there are no visual speech cues available. We speculate that this interpretation could be valid in the present study because most participants had good visual acuity. Nevertheless, we recognize that there is a distinction between structure and function and that the association between neural activation and cortical thickness is not straightforward.

As mentioned above, we only found very wide-spread and negative associations between CDTT SRTs and brain structure in the AD group who typically have more pronounced brain atrophy than the SCD or MCI groups. In the other two groups, we found few and only very focal positive associations between CDTT SRTs and cortical thickness (SCD: left posterior cingulum, right superior temporal gyrus; MCI: right superior frontal gyrus, right lateral orbitofrontal gyrus, and anterior cingulum). Given that participants in these groups likely have less brain atrophy than participants in the AD group, our findings may indicated that individuals who have more structural integrity in brain regions involved in cognitive processes (Metzler-Baddeley et al., 2012; Schermuly et al., 2010) and during speech-in-noise perception (Bidelman and Howell, 2016; Du et al., 2016) might be able to use these regions to compensate for difficulties understanding speech-in-noise (Rönnberg et al., 2013).

Overall, our data provide evidence that speech-in-noise recognition may be disproportionately impaired in older adults with AD. Our results further show that persons with AD who may have more structural decline in the frontal, temporal, and parietal lobes perform worse on the CDTT. It is possible that dementia-related neuropathology, which leads to accelerated decline of brain structures, may exacerbate or lead to a decline in speech processing in adverse listening conditions. Alternatively, declines in auditory processing may have wide-spread effects on brain atrophy in older adults with AD. Longitudinal studies are needed to clarify the direction of these effects.

### 4.3. Limitations

One limitation of the present study is that standard pure-tone audiometric thresholds were not obtained in COMPASS-ND because of the time-efficient audiometric screening protocol that was used. The resulting pure-tone hearing loss categories pose some challenges for parametric statistical analyses because of their categorical nature and do not allow the computation of conventional pure-tone averages which are typically used in similar studies. However, we were able to validate the hearing loss categories by demonstrating that they mapped to grades of hearing determined based on full audiograms in two independent samples of older adults. Additionally, we have limited information about the hearing aid usage or the length (e.g., age of onset) or nature (e.g., etiology) of hearing loss. A second limitation of the present study is the risk of Type I error given the large number of correlations that were examined. In our FreeSurfer analysis, we used an adjusted *p*-value (*p*<.001) to correct for multiple comparisons as has been done in previous research in which cortical thickness was examined as a function of hearing-related disorders such as Tinnitus (Meyer et al., 2016; Vanneste et al., 2015). A family-wise error correction or similar approaches such as Bonferroni correction would have been more rigorous in controlling Type I error; however, some epidemiologists and biostatisticians argue that these approaches might be too conservative (Perneger, 1998; VanderWeele and Mathur, 2019). The correlations conducted between the hearing measures and hippocampal volume did not survive Bonferroni correction, while the correlations between hearing loss category and CDTT SRTs do. We therefore recommend using larger datasets, for example with more data coming from COMPASS-ND in the future, for this type of research in the future, even though it is extremely challenging to recruit larger samples in this research.

A third shortcoming is the cross-sectional nature of the study. In order to clarify the direction of the associations between hearing loss and brain volume/thickness, longitudinal data are needed. In the COMPASS-ND study, longitudinal data are currently being collected from the same participants. Thus, the current report is a first step toward understanding the possible effects of two measures of hearing loss on brain structures in three selected diagnostic groups.

Finally, our findings may not generalize to individuals with SCD, MCI, and AD in the general population. Currently, our diagnostic groups have skewed sex ratios, which may not be representative of the diagnostic groups. Also, our participants were self-selected volunteers who agreed to be in a research study involving the administration of an extensive test battery.

### 4.4. Conclusion

Our study investigated the association between two measures of hearing loss and brain structure in older adults with or at risk for dementia across three diagnostic groups (SCD, MCI, AD). In those older adults with cognitive complaints only (SCD), those with greater pure-tone hearing loss had lower hippocampal volumes (a biomarker of dementia) and greater cortical thickness in auditory and higher-order language-related areas, potentially reflecting the effects of compensation. No such associations between pure-tone hearing loss category and brain structure were found in the MCI or AD groups. Taken together, we suggest that hearing loss is associated with reductions in hippocampal volume early in the dementia-risk spectrum and that AD-related pathological atrophy comes obscure the neuroanatomical effects of pure-tone hearing loss in later disease stages. Confirmation of this hypothesis awaits longitudinal data. Nevertheless, in the AD group only, we found that those with greater (worse) CDTT SRTs exhibited lower cortical thickness globally in both hemispheres. It is therefore possible that an age-related decline in speech-in-noise understanding may result in declines in brain structures in older adults who have (mild) cognitive impairment. Alternatively, and because the AD group performed slightly worse in the speech-in-noise task compared to the SCD group, it is also possible that dementia-related atrophy in brain regions supporting auditory processing leads to a decline in speech-in-noise understanding. Longitudinal research is needed to clarify the potential causal mechanisms. Our work extends the research on hearing-brain associations to include those who have subjective or objective cognitive complaints and suggests that these relationships evolve as disease progresses.

## Data Availability

Data information on: https://ccna-ccnv.ca/compass-nd-study/

## Acknowledgments

This research was supported by an infrastructure and operating grant to the CCNA from the Canadian Institutes of Health Research (CIHR) (Grant no. CNA-137794). This grant supports the COMPASS-ND study and the work of CCNA Team 17 principal investigators (NP, KPF, PM, JBO, WW) and trainees (NG, FA, SR). NG was supported by an Early Postdoc Mobility grant from the Swiss National Science Foundation (grant nr. P2ZHP1_174865). We gratefully acknowledge the important contributions of the COMPASS-ND PIT team, especially Victor Whitehead. We are grateful to Samantha Bishundayal and the Phillips CAP Lab for their contributions. We thank the COMPASS-ND research participants and site staff for their time. The data for the COMPASS-ND hearing loss validation was made possible using the data collected by the Canadian Longitudinal Study on Aging (CLSA). Funding for the CLSA is provided by the Government of Canada through the CIHR under grant reference: LSA 94473 and the Canada Foundation for Innovation. Part of this research has been conducted using the CLSA Baseline Comprehensive dataset version 3.2, under Application Number 160605. The CLSA is led by Drs. Parminder Raina, Christina Wolfson, and Susan Kirkland. The opinions expressed in this manuscript are the authors’ own and do not reflect the views of the Canadian Longitudinal Study on Aging.

## Footnotes

1 https://www.who.int/mental_health/neurology/dementia/guidelines_risk_reduction/en/

2 Based on a X^2^ test, there were no differences in the number of males and females in HL category in SCI, MCI, or AD.

3 Running an ANOVA with sex and diagnostic group as between-subjects variables, we did not find any differences between males and females in the SRTs on the CDTT in SCI, MCI, or AD.

## Notes

### Competing Interest Statement

The authors have declared no competing interest.

### Author Declarations

The COMPASS-ND study was approved by the Jewish General Hospital Research Ethics Board.

